# Food Insecurity as a Moderator of Rural Mental Health: A County-Level Analysis

**DOI:** 10.64898/2026.08.25.26361353

**Authors:** Eashwar Krishna, Neha Shanavas, Parthav Gavini, Cristy Roso

## Abstract

**Objective:** To examine if food insecurity moderates the relationship between rurality and mental health outcomes (suicide mortality, poor mental health days, frequent mental distress) and to assess if these effects vary across U.S. Census divisions.

**Methods:** This county-level (n=2,397) cross-sectional study used OLS and spatial error regression to analyze public data from sources including the County Health Rankings and USDA. We modeled suicide mortality, poor mental health days, and frequent mental distress as functions of the Index of Relative Rurality (IRR) and food insecurity, controlling for median income and provider rates. The suicide model was also tested across nine U.S. Census divisions.

**Results:** Baseline models revealed a paradox: rurality was a direct risk factor for suicide (B=0.400) but protective for poor mental health days (B=-0.224). The national multivariable model revealed a significant, positive rurality-food insecurity interaction for suicide mortality (B=0.861), indicating a synergistic risk. This interaction was not significant for general mental distress, which was more strongly predicted by income and food insecurity. Regional analysis confirmed the suicide interaction was potent in five divisions, including the Pacific (B=3.048) and Mountain (B=1.712), but absent in others (e.g., South Atlantic).

**Conclusions:** The drivers of suicide are distinct from those of general mental distress and are geographically heterogeneous. The interaction of rurality and food insecurity creates a compounded risk for suicide. Suicide prevention must be regionally-tailored and address structural inequalities, such as food insecurity, alongside clinical care.

## Introduction

Rural suicide rates in the United States are consistently and significantly higher than those in urban areas. This disparity is especially pronounced among men, with suicide rates increasing along a gradient of increasing rurality (Pettrone and Curtin, 2020). Moreover, the rural-urban suicide gap has widened over time, suggesting that rural communities are facing escalating socioeconomic pressures (Fontanella et al., 2015; Ivey-Stephenson et al., 2017).

A growing body of global literature has linked suicidal ideation with severe food insecurity across diverse demographic groups (Koyanagi et al., 2019). These effects are not uniform across populations; food insecurity has particularly strong mental health effects among vulnerable groups (Chai, 2024; Hasan et al., 2024), such as Indigenous populations in North America who already face elevated suicide risk (Ivey-Stephenson et al., 2017). Adverse mental health patterns in relation to food insecurity are well-documented among children and adolescents, but have been shown to be valid across international age groups (Azupogo et al., 2023; Thielman et al., 2023; Umutoniwase et al., 2022; Rahi et al., 2025). However, the literature on the U.S. remains relatively limited in scope, often focusing narrowly on young adults rather than broader age distributions or minority groups. Among these younger cohorts, the evidence is consistent: food-insecure young adults are more likely to experience depression, anxiety, and suicidal ideation (Nagata et al., 2019), and food-insecure college students are more likely to engage in self-injurious behaviors (Oh et al., 2022). In broader population samples, food insecurity has been associated with increased incidence of moderate to severe psychological distress (McClellan and Zuvekas, 2024).

In addition to structural factors, cultural and attitudinal barriers prevalent in rural communities – such as stoicism, stigma surrounding mental illness, and distrust of formal institutions – are likely to contribute to both the underreporting of psychological distress and the elevated rates of suicide mortality observed in these areas. These attitudinal features may influence not only whether individuals seek help, but also how mental health outcomes manifest across different levels of severity. Such dynamics are unlikely to be uniform across the country, as they may vary by region and intersect with demographic characteristics (eg. educational attainment) warranting a closer examination of inter-regional heterogeneity in observed patterns.

This study is grounded in research that specifies food insecurity and rurality shape suicide rates. Informed by insights from Durkheim’s concept of anomie, a form of normlessness and disorientation resulting from rapidly changing social structures, the present study examines the role of food insecurity in moderating the link between rurality and suicide in rural America, and if this role may vary across regions.

### Theoretical Framework

Emile Durkheim’s foundational sociological theory of suicide posits that its causes lie not solely within the individual, but in the structure of social relationships, particularly in two domains: social integration and social regulation (Durkheim, 2005). Social integration refers to the degree of attachment and sense of belonging individuals feel toward their community; a deficit in integration can produce egoistic suicide, in which disconnected individuals perceive life as meaningless. Social regulation, by contrast, involves the external norms that shape and guide behavior. Its breakdown results in anomic suicide where individuals experience moral disorientation due to the absence of predictable norms or institutional guidance. This type becomes prevalent during rapid social or economic change, disrupting habitual expectations, practices, and identities.

Recent elaborations on Durkheim’s theory have emphasized the socioemotional underpinnings of suicide, particularly the role of shame in mediating how individuals experience regulatory breakdowns. As Abrutyn and Mueller (2014) argue, regulation is not only institutional but also emotional: it provides individuals with cultural clarity and moral expectations that help stabilize identity. When these expectations collapse, such as during economic dislocation or cultural displacement, individuals may encounter not just confusion but a profound sense of violated selfhood. The emotional core of anomic suicide, then, is not sadness alone, but a volatile mixture of shame, frustration, and weariness. This form of shame arises when one’s social roles or expectations are disrupted, particularly when those roles are central to one’s self-concept. It is often triggered by sudden threats to status, identity, or perceived social value. Unlike egoistic forms of despair, anomic shame may remain repressed or bypassed until it culminates in a breaking point: sometimes expressed through impulsive or fatal acts. In this view, suicide does not result solely from external dislocation but from an internalized crisis of identity, a painful realization of having become misaligned or excluded from the very moral order that once gave life meaning.

The relevance of Durkheim’s framework to the American rural suicide crisis is, in our eyes, increasingly evident. Rural areas exhibit disproportionately high rates of “deaths of despair” including suicide, drug overdoses, and alcohol-related mortality (Lee et al., 2024). We argue that rising rural suicide rates can be partially explained through a Durkheimian lens: the breakdown of rural economic structures and the erosion of traditional identities has produced conditions of anomie. In rural communities, the commercialization of agriculture has destabilized local food systems, driven up the costs of essential agricultural equipment, displaced small family farms, and disrupted generational identities (Lobao and Stofferahn, 2007; Lacy, 2018). The widespread decline of small family farms has been well-documented (Robbins, 2023; Clark, 2024), and these farms have been essential to rural identity and social cohesion (Munch, 2024). These processes may not only increase material deprivation but also fracture social cohesion and diminish perceived agency over food access and community wellbeing, reflected in organized responses to cultural and economic displacement (Van Dyke & Soule, 2002). Consequently, food insecurity in rural areas may act not merely as a nutritional or economic stressor but as a environmental condition that behaves as an intensifier of anomic conditions that elevate suicide risk.

While the independent associations between rurality and suicide, and between food insecurity and mental health outcomes, are well-established, their interaction remains unexplored. To date, few, if any, national-level quantitative studies have tested whether food insecurity intensifies the relationship between rurality and suicide mortality, leaving a critical gap in the literature. This oversight persists despite theoretical and qualitative evidence suggesting that material hardship and social destabilization interact in ways that could heighten mental health outcome risk.

This study addresses these gaps by empirically evaluating the individual and interactive effects of rurality and food insecurity on suicide mortality and psychological distress. It also examines whether these relationships are consistent across the United States or exhibit significant regional variation. In doing so, the present work brings a classical sociological framework into dialogue with contemporary epidemiological methods, offering a novel account of the structural and cultural forces driving rural mental health disparities.

## Methods

### Data Sources

This study draws on publicly available county-level data for the United States. The primary predictors and outcome variables were obtained from the 2025 County Health Rankings dataset, which compiles data from national surveillance sources including the National Center for Health Statistics (NCHS) and the Behavioral Risk Factor Surveillance System (BRFSS) (Population Health Institute, 2025). Rurality was measured using the 2020 Index of Relative Rurality (IRR), a continuous metric that integrates population size, density, remoteness, and built environment (Ayoung and Waldorf, 2023). Regional divisions followed the standard classification schema of the U.S. Census Bureau. County-level median household income data were retrieved from the U.S. Department of Agriculture’s Economic Research Service (USDA, 2025b).

### Statistical Analysis

The final analytic sample consisted of (n = 2,397) U.S. counties with complete data across all variables of interest. Data cleaning involved a combination of manual exclusion and automated listwise deletion in R. All variables, including predictors, outcomes, and controls, were min-max normalized to a continuous 0-1 scale to allow for interpretable and comparable regression coefficients across models. For the IRR, which is originally expressed on a 0-1 scale, min-max normalization was still performed post-filtering to ensure consistency with the final sample’s value distribution. In this normalized scale, a value of 0 corresponds to the least rural (most urban) county and a value of 1 to the most rural.

Rurality was operationalized using the Index of Relative Rurality (IRR), a multidimensional metric designed to move beyond binary rural-urban categorizations by incorporating multiple dimensions of rurality including population size, density, and remoteness (Ayoung and Kim, 2023). Food insecurity was measured using the percentage of the population experiencing food insecurity at the county level, sourced from County Health Rankings based on data provided by Feeding America’s Map the Meal Gap project (Population Health Institute, 2025). Food insecurity was defined using the USDA’s Core Food Security Module (CFSM), with households classified as insecure if they answered affirmatively to at least three items from the module (Feeding America, 2024). The CFSM has been previously validated in both national and subpopulation samples (USDA, 2025a; Frongillo, 1999).

Mental health outcomes were assessed using three measures, each sourced from BRFSS or NCHS data as disseminated through County Health Rankings. Suicide mortality was measured as the age-adjusted rate of suicide deaths per 100,000 residents. Frequent mental distress was defined as the percentage of adults reporting symptoms of stress, depression, and other states on 14 or more days in the past 30. The average number of poor mental health days, a continuous variable, captured the mean number of days within a 30-day period that respondents reported experiencing poor mental health. These three outcomes were chosen to capture both severe and subclinical dimensions of psychological strain, aligning with the study’s emphasis on food insecurity as both a material stressor and a sociological destabilizer. Two covariates were included in all models to adjust for potential confounding: median household income, sourced from the USDA (USDA, 2025b), and the county-level rate of mental health providers per 100,000 population, derived by County Health Rankings from the Centers for Medicare and Medicaid Services’ National Provider Identifier file (Population Health Institute, 2025).

We employed Ordinary Least Squares (OLS) regression to estimate associations between county-level rurality, food insecurity, and mental health outcomes. To account for the nested data structure, all models incorporated robust standard errors clustered at the state level. The analytical strategy proceeded in multiple phases. First, bivariate OLS regressions assessed the independent effects of rurality and food insecurity on suicide mortality, poor mental health days, and frequent distress, while controlling for county-level median income and mental health provider rate. A separate hierarchical regression was conducted to evaluate whether the association between rurality and food insecurity was attenuated by income, establishing the structural basis for interaction modeling.

Next, multivariable regressions introduced a rurality-food insecurity interaction term to assess whether the effect of rurality on mental health outcomes was moderated by food insecurity. Fully adjusted models included both predictors, the interaction term, and the two covariates. To assess geographic heterogeneity, we estimated the full suicide mortality model separately across the nine U.S. Census divisions. Multiple post-hoc analyses were conducted to evaluate the stability of the primary suicide mortality model; see Appendix B for details. All regression models were implemented in R (Version 4.4.1) using RStudio (Version 2025.05.1+513), and independently replicated by both authors.

## Results

In the initial regression models assessing the independent effects of rurality and food insecurity on mental health outcomes, rurality was a statistically significant predictor of suicide mortality. Higher levels of rurality were associated with elevated suicide rates (B = 0.400, p < 0.001). In contrast, rurality was negatively associated with average poor mental health days (B = –0.224, p < 0.01), while its inverse association with frequent mental distress did not reach statistical significance (B = –0.148, p > 0.05).

Food insecurity, when entered independently, was not significantly associated with suicide mortality (B = 0.063, p > 0.05). However, it was strongly associated with worse general mental health outcomes, including more poor mental health days (B = 0.389, p < 0.001) and higher rates of frequent distress (B = 0.484, p < 0.001).

Across models, county-level median household income exhibited a consistent and protective association. Higher income was significantly related to fewer poor mental health days (B = –0.670, p < 0.001), lower rates of frequent distress (B = –0.888, p < 0.001), and modestly lower suicide mortality (B = –0.051, p > 0.05 in the rurality model; B = –0.133, p < 0.01 in the food insecurity model). The mental health provider rate was positively associated with suicide mortality in the rurality model (B = 0.179, p < 0.01), but showed no significant association with the distress measures.

To further clarify the structural determinants of food insecurity, a supplementary regression was estimated using food insecurity as the dependent variable. In the unadjusted model, rurality was positively associated with food insecurity (B = 0.257, p < 0.001). However, this association reversed when controlling for median income (B = –0.130, p < 0.001), which emerged as the dominant predictor (B = –0.987, p < 0.001, R² = 0.537). This reversal suggests that the apparent rural disadvantage in food access is largely attributable to lower income levels in rural counties.

In the fully adjusted interaction models including rurality, food insecurity, their multiplicative interaction term, and both covariates, the relationship between predictors and outcomes diverged depending on the mental health outcome (Table 2). To address spatial autocorrelation in the residuals of the initial OLS models (Moran’s I = 0.29, p < 0.001), spatial error models with state-level fixed effects were estimated for each mental health outcome. The inclusion of a spatial error term reduced residual spatial autocorrelation substantially (Moran’s I = –0.0079, p = 0.710) and improved model fit, as reflected in lower AIC values across all outcomes.

**Table 1.** Baseline Regression Effects.

| *Variable* | **IRR vs.**<br>**Suicide** | **IRR vs.**<br>**Poor Days** | **IRR vs.**<br>**Distress** | **FI vs.**<br>**Suicide** | **FI vs.**<br>**Poor Days** | **FI vs.**<br>**Distress** |
| --- | --- | --- | --- | --- | --- | --- |
| Rurality<br>(IRR) | 0.400*** | -0.224** | -0.148 | -- | -- | -- |
| Food<br>Insecurity<br>(FI) | -- | -- | -- | 0.063 | 0.389*** | 0.484*** |
| Median<br>Income | -0.051 | -0.670*** | -0.888*** | -0.133** | -0.226** | -0.382*** |
| Provider<br>Rate | 0.179** | 0.043 | -0.061 | 0.057 | 0.064 | -0.078 |

*Standardized regression coefficients (B) reported. \*\*\* $p < 0.001$ , \*\* $p < 0.01$ , \* $p < 0.05$*

**Table 2.** Multivariate Regression Assessing the Impact of Rurality and Food Insecurity on Mental Health Outcomes.

|  | **Suicide**<br>**Mortality** | **Poor Mental**<br>**Health Days** | **Frequent Mental**<br>**Distress** |
| --- | --- | --- | --- |
| *Predictors* |  |  |  |
| Rurality | -0.056 | 0.020 | 0.102 |
| Food Insecurity (FI) | -0.351*** | 0.212*** | 0.229*** |
| Rurality × FI | 0.861*** | 0.145 . | 0.317** |
| *Controls* |  |  |  |
| Median Income | -0.075*** | -0.303*** | -0.372*** |
| Provider Rate | 0.029 | -0.012 | -0.102*** |

*Table displays coefficients (B) from Spatial Error Models with state-level fixed effects. Significance Levels: . $p < 0.10$ , \* $p < 0.05$ , \*\* $p < 0.01$ , \*\*\* $p < 0.001$ .*

**Table 3.** Multivariable Regressions Assessing the Impact of Food Insecurity and Rurality on Suicide Mortality by Census Division.

|  | **Rurality** | **Food Insecurity** | **Interaction** | **Median Income** | **Provider Rate** | **R<sup>2</sup>** |
| --- | --- | --- | --- | --- | --- | --- |
| East North Central | -0.149 | -0.438 | **1.030\*** | -0.163 | -0.043 | 0.348 |
| East South Central | 0.332 | 0.082 | -0.176 | 0.041 | -0.021 | 0.093 |
| Mid-Atlantic | 0.041 | -0.156 | 0.424 | -0.113 | -0.016 | 0.525 |
| Mountain | -0.172 | -0.926* | **1.712\*** | -0.174 | -0.111 | 0.334 |
| New England | 0.179** | -0.076 | **0.474\*\*** | 0.005 | -0.201* | 0.781 |
| Pacific | -0.554** | -1.620** | **3.048\*\*\*** | -0.270* | 0.104 | 0.829 |
| South Atlantic | 0.191** | 0.008 | -0.051 | -0.069 | 0.029 | 0.188 |
| West North Central | -0.113 | -0.655 | 1.479 | -0.086 | -0.133* | 0.367 |
| West South Central | -0.015 | -0.403 | **0.848\*** | 0.003 | 0.235 | 0.245 |

*Table displays coefficients (B) from Spatial Error Models with state-level fixed effects.*
*Significance Levels: . $p < 0.10$ , \* $p < 0.05$ , \*\* $p < 0.01$ , \*\*\* $p < 0.001$ .*

In the final spatial error model predicting suicide mortality, the interaction between rurality and food insecurity remained statistically significant (B = 0.861, p < 0.001), suggesting a robust compounding association between these two structural factors. Food insecurity was negatively associated with suicide mortality in the main effect (B = –0.351, p < 0.001), while rurality alone showed no significant direct effect (B = –0.056, p > 0.05) while controlling for the other variables. Median income retained a modest protective association (B = –0.075, p < 0.001), while the provider rate showed no significant relationship (B = 0.029, p > 0.05). The estimated spatial error parameter (λ = 0.219, p < 0.001) indicates residual spatial dependence even after controlling for state effects, justifying the use of spatial modeling.

In the model predicting average poor mental health days, food insecurity was positively associated with the outcome (B = 0.212, p < 0.001), and the rurality-food insecurity interaction reached marginal significance (B = 0.145, p < 0.10). Median income again exhibited a strong negative association (B = –0.303, p < 0.001), while neither rurality nor provider rate were significant. The spatial error parameter for this model was λ = 0.289 (p < 0.001).

For frequent mental distress, food insecurity remained a significant positive predictor (B = 0.229, p < 0.001), and the rurality-food insecurity interaction was also significant (B = 0.317, p < 0.01). Median income was again negatively associated with the outcome (B = –0.372, p < 0.001), while rurality showed no significant main effect (B = 0.102, p > 0.05) having controlled for the other effects. Provider availability was inversely associated with frequent distress (B = –0.102, p < 0.001), and the spatial error parameter remained significant (λ = 0.234, p < 0.001).

The spatial error models with state-level fixed effects revealed significant geographic heterogeneity in mental health outcomes; see Appendix A for state-level findings.

To assess geographic heterogeneity in the relationship between rurality, food insecurity, and suicide mortality, fully adjusted models including main effects, an interaction term, and covariates were estimated separately for each of the nine U.S. Census divisions using robust standard error clustering at the state level. The results revealed substantial variation in both the magnitude and statistical significance of the predictors across regions.

The interaction between rurality and food insecurity was significantly and positively associated with suicide mortality in five of the nine divisions. The Pacific division exhibited the largest interaction coefficient (B = 3.048, p < 0.001), followed by the Mountain division (B = 1.712, p < 0.05). A similar pattern was observed in the West South Central division, where the interaction term was also significant (B = 0.848, p < 0.05). In the East North Central division, which includes much of the industrial Midwest, the interaction coefficient was also positive and significant (B = 1.030, p < 0.05). New England likewise exhibited a significant interaction effect (B = 0.474, p < 0.01), though the magnitude was smaller than in Western divisions.

In contrast, the interaction term did not reach statistical significance in the South Atlantic, East South Central, Mid-Atlantic, or West North Central divisions. In these areas, no consistent pattern emerged in the main effects of rurality or food insecurity on suicide mortality. For example, in the South Atlantic and East South Central divisions, the coefficients for both rurality and food insecurity were not statistically significant, and the interaction terms were close to zero (B = −0.051 and B = −0.176, respectively). Similarly, the Mid-Atlantic division showed modest and nonsignificant main and interaction effects (B = 0.424).

The explanatory strength of the models, as indicated by R² values, also varied substantially. The highest model fit was observed in the Pacific division (R² = 0.829), followed by New England (R² = 0.781), and the Mid-Atlantic (R² = 0.525), albeit the latter model’s statistical insufficiency. In contrast, divisions such as the South Atlantic (R² = 0.188) and East South Central (R² = 0.093) exhibited relatively low explanatory power, suggesting greater variability or unmeasured factors influencing suicide mortality in these regions. The results of the robustness analyses were strong and supported the stability of the full model; see Appendix B for specific results.

## Discussion

This study aimed to disentangle the complex effects of rurality and food insecurity on different mental health outcomes, with a specific focus on suicide mortality and mental distress. Three major findings were revealed that highlighted the intricate relationship between food insecurity, mental health, and geography. First, we identified a compelling “rural paradox”: while rurality emerged as a direct and statistically significant risk factor for suicide mortality, it was paradoxically associated with better general mental health outcomes, including fewer reported poor mental health days. This seemingly contradictory finding could be a result of systematic underreporting of mental health struggles in rural areas, potentially due to cultural stigma or limited mental health awareness (Cheesemond et al., 2019; Yang et al., 2011).

Second, our analysis revealed a unique synergistic interaction between rurality and food insecurity in predicting suicide mortality. Within categories of rurality, food insecurity consistently associated with higher suicide mortality, and when comparing food insecure urban and rural counties, rural areas demonstrated substantially higher suicide rates. Our Johnson-Neyman analysis revealed critical thresholds: once a county’s rurality score surpasses 0.52 on the normalized scale, food insecurity becomes a statistically significant risk factor for higher suicide mortality (Kim & Waldorf, 2023). Third, we found that this harmful interaction displays significant regional heterogeneity even after accounting for substantial spatial autocorrelation through state level fixed effects. While we were able to resolve 57% of spatial autocorrelation through state fixed effects, residual spillovers indicate unmeasured cross border dynamics that future research should address.

The conflicting roles of rurality in our findings reveal how geographical context shapes mental health outcomes differently across the severity spectrum. The literature is mixed regarding a ’protective effect’ for rural residents. While some studies suggest rural life is associated with stronger social relationships (e.g., Pérès et al., 2021), this finding does not reliably translate to a consistent mental health advantage. In fact, other research has identified significant or even higher rates of loneliness among rural populations (Abshire et al., 2020). This complexity becomes particularly critical when looking at severe, potentially life-threatening conditions. For these conditions, other aspects of rural life can transform into significant barriers. Contemporary research has identified enduring attitudinal barriers such as stoicism and institutional distrust in rural populations that diminish help-seeking tendency. Critically, among individuals with severe mental illness, research shows that the sense of community connection is actually stronger in urban rather than rural areas (Cheesemond et al., 2019).

Food insecurity operates as a powerful, multifaceted stressor through both physiological and psychological pathways. From a physiological perspective, proper nutrient intake correlates with positive mental health outcomes, while high–glycemic index dietary patterns have been linked to higher risk of incident depression (Gangwisch et al., 2015; Firth et al., 2020). However, the psychological burden may be more relevant to suicide risk. The consistent worry about food quantities represents a major stressor, and the inability to provide adequately can cause feelings of shame and profound embarrassment (Lobao & Stofferahn, 2007). Most critically, the lack of choice and agency over one’s diet can generate profound frustration and reduced sense of control -- both well-established risk factors for suicidal behavior.

The relationship between food insecurity, rurality, and suicide mortality may represent a particular kind of deprivation: a loss of traditional communal control over food systems amidst general economic distress. The dramatic transformation from small family farms to large-scale corporate agriculture has contributed to rural anomie, characterized by pervasive normlessness and social disorientation. Within this anomic context, food insecurity transcends simple material deprivation and becomes a reflection of identity disintegration (Goldschmidt 1978; Durkheim, 1952). The lived experience of food insecurity in areas historically thought of as the country’s “breadbasket” produces psychological distress that is clearly more severe than the sum of its parts, aligning with Durkheim’s theory on anomic suicide (Liebe et al., 2025; Gearing et al., 2021). In rural settings where self-sufficiency and agricultural identity are central, food insecurity may be experienced as disempowerment, symbolizing broader community decline and cultural displacement. This is especially true in multigenerational farming families where the farm is more than a livelihood: it is a legacy, an identity, and a moral commitment (Batey et al., 2023). Many farmers who died by suicide described a collapse of their ability to fulfill the ’farmer’ role -- compounded by what has been termed a “community panopticon” -- a form of social surveillance that intensified their distress (Purc-Stephenson et al., 2023). Our findings reveal striking geographic heterogeneity in how rurality and food insecurity interact to influence suicide mortality. The Pacific region demonstrated the most pronounced interaction effects, which could be due to areas with deeply rooted farming traditions experiencing severe anomic disruption when agricultural systems undergo change (Durkheim, 1952). In contrast, the South Atlantic and East South Central regions showed absence of significant interaction effects, possibly signaling regional resilience through continuation of traditional farming practices or distinct socioeconomic structures. The substantial regional variation constitutes a significant finding that calls for additional geography-based research to reveal the unique cultural and structural contexts that shape suicide risk across different rural American communities.

### Policy and Public Health Implications

This study’s findings offer important insights for policymakers and public health practitioners who are trying to address the growing crisis of suicide mortality in rural America. The statistically significant interaction between rurality and food insecurity shows that suicide risk is shaped by the social and structural contexts. Rurality magnifies the psychological and sociological effects of food insecurity, particularly in regions undergoing agricultural, economic, and cultural upheaval. These results support the argument that suicide prevention must be approached as both a mental health and structural inequality issue, rather than as a matter of individual risk alone. To fully understand this crisis, it is crucial to pay attention to the broader context of rural structural violence. Decades of economic disinvestment, agricultural consolidation, and policy neglect have systematically eroded community infrastructures and intensified psychosocial distress (Goldschmidt 1978; Durkheim, 1952). In this light, suicide in rural areas may be better understood not as isolated challenges but rather the consequence of systemic abandonment and enduring injustice.

Given these psychosocial dimensions, it follows that policies aimed at bolstering local food systems could have far-reaching benefits beyond nutrition alone. Policies that support local food infrastructure, such as cooperative grocery stores, community service agriculture (CSA), and small-scale food retailers, can play a crucial role in restoring both material access and a sense of agency. Programs like the Michigan Good Food Fund serve as promising models for rebuilding food sovereignty while also enhancing social cohesion (Michigan Good Food Fund, 2025). For example, expanding mobile grocery access not only improves nutrition but also reduces isolation and may be associated with lower depressive symptoms -- both critical protective factors against suicide (Nagata et al., 2019). Similarly, behavioral health integration in agricultural extension programs embeds support in trusted community institutions, circumventing traditional barriers of stigma and access.. Additionally, federal nutrition programs like SNAP and WIC, though widely used in rural areas, are often underutilized due to stigma or logistical barriers (Gearing et al., 2021; Children’s HealthWatch, 2013). Policymakers should prioritize expanding access through mobile food distribution, increased program visibility, and community-based outreach. Programs like the Geisinger Fresh Food Farmacy in rural Pennsylvania illustrate how health systems can play a direct role in addressing upstream social determinants of suicide risk. Geisinger’s model allows physicians to ’prescribe’ fresh, healthy food to food-insecure patients, who then receive groceries and nutritional education at clinic-based Farmacies (Fresh Food Farmacy, 2025).

Beyond food access, the severe shortage of mental health providers in rural America remains a fundamental structural barrier that compounds the crisis. Over 122 million Americans live in areas designated as mental health professional shortage areas, and rural communities are disproportionately affected (Lennartz, 2025). However, our adjusted model findings suggest that increasing provider density alone is not sufficient to reduce suicide mortality. While workforce expansion is necessary, it must be accompanied by measures that address affordability, transportation, institutional distrust, and stigma. In this regard, expanding telehealth services -- supported by clear licensure pathways (e.g., the Interstate Medical Licensure Compact) and reimbursement at parity with in-person care -- offers a promising approach (Physician License |

Interstate Medical Licensure Compact, 2025). Evidence from rural veterans and other populations suggests that telemental health can be as effective as in-person care for many outcomes (Watanabe et al., 2023). States should also invest in targeted recruitment and retention strategies, such as loan repayment programs and rural clinical training pipelines. West Virginia’s Rural Health Initiative provides one example of a long-term investment in building local capacity (West Virginia Resources - Rural Health Information Hub, n.d.). Furthermore, policies should support the use of peer support specialists and non-clinical providers, as well as Mental Health First Aid training within rural communities, to increase early intervention and culturally grounded care.

Cultural norms around stoicism and self-reliance, which is seen commonly in the rural Midwest and Appalachia, present unique challenges to traditional public health interventions. Our findings echo prior research showing a disconnect between high suicide mortality and low self-reported mental distress in places like West Virginia and the Plains states (Davis et al., 2024). This is reinforced by qualitative data from Purc-Stephenson et al., which found that men who died by suicide were often described as hardworking and emotionally guarded -- shaped by hegemonic ideals of masculinity and perceived community surveillance, what they term the “community panopticon”. In such contexts, psychological help-seeking may be seen as a personal failure, compounding isolation and distress. Addressing these cultural barriers requires public health efforts to invest in qualitative research, including ethnographic methods and in-depth interviews, that better capture the lived experiences of distress in culturally distinct rural communities. These approaches are especially important in regions where survey data may fail to capture the true scope of suffering. Community-led needs assessments and planning efforts should also be supported through long-term funding. This should be supplemented by mental health professionals who must receive training in cultural humility and contextual competence.

Incorporating traditional healing practices, faith-based support, and multilingual services into care delivery can enhance trust and improve outcomes. Mental health messaging should also be reframed using language that aligns with local values; for example, terms like “resilience,” “stress management,” or “wellness” may resonate more than clinical diagnoses. Campaigns like South Dakota’s rural anti-stigma initiative and outreach efforts such as the TransFARMation podcast demonstrate how rural-appropriate messaging can effectively engage hard-to-reach populations (The Transfarmation Project, 2025; Lurken & Lurken, 2023).

Our results call for coordinated, intersectoral policymaking that breaks down arbitrary barriers between agriculture, public health, economic development, and education. Suicide prevention in rural America requires a broad policy lens that incorporates food system resilience, workforce development, insurance reform, and economic stabilization. Policies that address rural food insecurity, improve Medicaid reimbursement rates, and expand crisis services like the 988 Lifeline must be integrated into broader rural health frameworks. States should apply a deliberate “rural lens” when evaluating new legislation, ensuring that policy decisions consider the geographic, cultural, and structural barriers unique to rural communities. The West Virginia State Rural Health Plan offers a model for how community, academic, and policy stakeholders can coordinate to address persistent disparities. Furthermore, we echo findings from recent agricultural resiliency research calling for targeted investments in technical assistance for farmers, strategic problem-solving support, and integrated behavioral health models in primary care (State Rural Health Plans - Rural Health Information Hub, n.d.; Purc-Stephenson et al., 2023).

### Strengths and Limitations

The utilization of a large, nationwide dataset solidifies the external validity of this work and allows broad generalizability across the U.S, likely even to counties that may have been excluded in the present analysis. A significant methodological advancement over conventional urban/rural binary classifications is also shown by the use of the Index of Relative Rurality (IRR), a continuous, multidimensional measure of rurality. This is crucial to our study as IRR better captures the complexity and variation across a spectrum of rurality, as compared to binary or ordinal classifications which necessarily group counties together that may or may not share relevant qualities (Kim & Waldorf, 2025). Durkheim’s theory of anomie explicitly grounds the study in traditional sociological theory. This gives our findings a solid theoretical foundation for comprehending intricate social processes. We also first established the baseline effects of rurality and food insecurity separately, then studied the interaction, which produced results that are more readily interpretable and actionable (Durkheim, 1952).

Despite several strengths, several inherent limitations warrant consideration. First, because our data are aggregated at the county level, the analysis can be susceptible to ecological fallacy; conclusions cannot be definitively drawn about specific individuals based on county-level associations. Second, the cross sectional nature of our data inhibits temporal understanding. This prevents definitive causal inferences from the observed relationships. While our theoretical framework suggests a causal direction, longitudinal studies are needed for establishing temporal precedence and strengthening causal claims. Third, it may be the case that suicide mortality is a narrow measure of anomie-induced death, and a broader outcome including all deaths of despair may more effectively capture the effect studied. Fourth, a non-negligible number of counties were excluded due to incomplete or missing data, and it is possible that counties that did not report data were more rural and disadvantaged than included counties. U.S territories were excluded as well. Finally, despite including key covariates such as median household income and mental health provider rates, the study may be subject to omitted variable bias. Other unmeasured contextual factors, like regional substance use rates, access to lethal means, specific community-level social capital metrics, could influence the observed outcomes and potentially confound the relationships we identified. The residual spatial dependence observed even after controlling for state fixed effects indicates that unmeasured cross-border dynamics may be operative, strongly suggesting that future research should incorporate mobility data and more sophisticated spatio-temporal modeling approaches.

### Future Directions and Vulnerable Populations

While our study examined food insecurity as a structural amplifier of suicide risk in rural America, further research is urgently needed to examine how this dynamic may affect vulnerable subpopulations. This includes racial/ethnic minorities, gender diverse individuals, Indigenous peoples, and veterans: groups who may experience additional layers of stigma, discrimination, or under-resourced care. Intersectional approaches and equity focused policy interventions must be prioritized in future studies. Models such as the rural telemental health for veterans are examples of starting points for designing more responsive care systems.

## Conclusion

In sum, this study provides strong evidence that suicide prevention in rural communities must go beyond clinical intervention and address the social, economic, and cultural conditions that shape vulnerability. Food insecurity, in particular, emerges not only as a material stressor but as a key sociological amplifier of risk -- especially in places where agricultural decline and loss of community identity are most acute. Preventing further loss will demand not only improved mental health services but deep investment in the social and economic conditions that underpin well-being in rural communities.

## Data Availability

All data produced in the present study are available upon reasonable request to the authors

https://www.countyhealthrankings.org/health-data/county-health-rankings-measures

## Acknowledgements

The authors would like to thank Dr. Ryan Talbert, Department of Sociology, University of Connecticut, for his guidance and support in the execution of this project.

## Related Presentations

This project was presented as a poster at the New England Rural Health Association conference on November 5th, 2025 in Groton, CT

